# MicroRNA-based cancer mortality risk scoring system and hTERT expression in early-stage oral squamous cell carcinoma

**DOI:** 10.1101/2020.07.03.20145854

**Authors:** Angela J. Yoon, Regina M. Santella, Shuang Wang, David I. Kutler, Richard D. Carvajal, Elizabeth Philipone, Tian Wang, Scott M. Peters, Claire R. Stewart, Fatemeh Momen-Heravi, Scott Troob, Matt Levin, Zohreh AkhavanAghdam, Austin J. Shackelford, Carleigh R. Canterbury, Masataka Shimonosono, Brenda Y. Hernandez, Bradley D. McDowell, Hiroshi Nakagawa

## Abstract

We have previously constructed a novel microRNA (miRNA)-based prognostic model and cancer-specific mortality risk score formula to predict survival outcome in oral squamous cell carcinoma (OSCC) patients who are already categorized into ‘early-stage’ by the TNM staging system. A total of 836 early-stage OSCC patients were assigned the mortality risk scores. We evaluated the efficacy of various treatment regimens in terms of survival benefit compared to surgery only in patients stratified into high (risk score ≥0) versus low (risk score <0) mortality risk categories. For the high-risk group, surgery with neck dissection significantly improved the 5-year survival to 75% from 46% with surgery only (p<0.001); a Cox proportional hazard model on time-to-death demonstrated a hazard ratio of 0.37 for surgery with neck dissection (95% CI: 0.2-0.6; p=0.0005). For the low-risk group, surgery only was the treatment of choice associated with 5-year survival benefit. Regardless of treatment selected, those with risk score ≥2 may benefit from additional therapy to prevent cancer relapse. We also identified hTERT (human telomerase reverse transcriptase) as a gene target common to the prognostic miRNAs. There was 22-fold increase in the hTERT expression level in patients with risk score ≥2 compared to healthy controls (p<0.0005). Overexpression of hTERT was also observed in the patient-derived OSCC organoid compared to that of normal organoid. The DNA cancer vaccine that targets hTERT expressing cells currently undergoing rigorous clinical evaluation for other tumors can be repurposed to prevent cancer recurrence in these high-risk early-stage oral cancer patients.

## INTRODUCTION

An estimated 49,000 people in United States are diagnosed with oral squamous cell carcinoma (OSCC) each year [1,2]. Leading etiologic factors include tobacco and alcohol [1-5]. While 80% of oropharyngeal cancers are related to high-risk human papillomavirus (HPV types 16 and 18), the incidence of high-risk HPV-related oral cancer is relatively low, estimated to be ∼4% [4,5]. Despite advances in cancer diagnosis and treatment, the overall 5-year survival rate for OSCC remains the lowest among malignancies and, in fact, has been <50% for the last three decades [1-5]. Among newly diagnosed oral cancer cases, ∼80% are in the tumor-node-metastasis (TNM) Stage I/II without regional lymph node involvement or distant metastasis [3]. Hence, a window of opportunity exists in which accurate prognostication and subsequent decisions for appropriate treatment could dramatically improve 5-year survival of patients with this deadly disease.

While the TNM stage is considered the key prognostic determinant in oral cancer [6], it is incapable of delineating individual risk for patients within the same TNM stage strata. To address these critical gaps, we have constructed a microRNA (miRNA)-based prognostic model and cancer-specific mortality risk score formula using a pool of early-stage oral cancer patients who had surgery only as initial treatment [7]. miRNAs are small, 18-24 nucleotide long, non-coding RNA molecules that regulate the expression of targeted genes either by facilitating mRNA degradation or by repressing translation [8,9]. One miRNA is capable of binding over 100 different mRNAs with different binding efficiencies and plays a crucial role in their post-transcriptional regulation [8-12]. miRNAs control cell growth, apoptosis and differentiation, and various types of cancer have demonstrated distinct miRNA expression profiles [8-12]. Thus far, a number of miRNAs associated with clinical outcomes have been reported for lung, breast, gastric and pancreatic cancers, as well as OSCC/head and neck carcinomas [12-14].

In our miRNA-based prognostic model, we included pertinent clinico-demographic covariates [7]. Therefore, the final prognostic model was built based on both expression of the miRNAs (miRNA-127-3p, 4736, 655-3p) and clinico-demographic covariates (TNM Stage I vs II, histologic grade well vs moderate/poor), which allowed for multiple risk factors to be used systematically and reproducibly to maximize the prognostic power. The prognostic model was subject to rigorous internal validation, followed by external validation, which included patient populations from Pennsylvania, Iowa and Hawaii [7]. The performance of the model, in terms of its discriminatory power to differentiate between high and low cancer-specific mortality risks, was meticulously verified. The area under the curve (AUC) of the receiver operating characteristic (ROC) curve with the miRNA-based 5-plex marker panel was 0.83 (p<0.001), 0.87 (p<0.001) and 0.81 (p<0.001) in internal test, internal validation and external validation cohorts, respectively, demonstrating uniformly significant prognostic value [7]. The clinical prognostic indicators alone, including TNM stage and histologic grading, had an AUC of 0.67 (p<0.001), demonstrating that the 5-plex prognostic marker significantly increase the prognostic power [7].

From the final prognostic model, we constructed a robust mortality risk score formula. This personalized formula consisted of the patient’s miRNA expression levels and prognostic covariates weighed by their regression coefficient. The main purpose of the risk score formula is for clinicians to easily translate miRNA levels obtained from the clinical laboratory, along with known clinico-demographic variables into the patient’s risk scores, which will serve as a practical method to assess patient-specific mortality risk in the clinical setting. The risk score from the 5-plex marker panel consisting of miRNAs-127-3p, 4736, 655-3p, TNM stage and histologic grading stratified patients initially into high (≥0) vs low (<0) risk categories, and then sub-risk stratified into finer risk categories [highest (risk score ≥2) vs moderately-high (risk score 1-2) vs moderately-low (risk score 0-1) vs low (risk score <0)]. Compared to the low-risk strata (<0), the highest-risk strata (≥2) had 23-fold increased mortality risk (hazard ratio of 23, 95% confidence interval 13-42), with a median time-to-recurrence of 6 months and time-to-death of 11 months (versus >60 months for both outcomes among low-risk patients; p<0.001).

In this study, we evaluated the efficacy of various treatment regimens in patients stratified into high vs. low mortality risk categories by the miRNA-based prognostic risk score formula. In specific, we assessed the median time-to-recurrence, the median time-to-death, and 5-year survival rate in patients who received surgery only (S) vs. surgery with neck dissection (S+ND) vs. surgery with irradiation (S+IR) vs. surgery with ND and IR (S+ND/IR) as the initial surgical treatment, and compared the survival benefit associated with each treatment.

## MATERIALS AND METHODS

### Subjects and Study Design

Following approval from the Institutional Review Board (IRB), 836 early-stage OSCC patients, ≥18 years old, newly-diagnosed with primary OSCC and with a minimum of 5-year clinical outcomes information were identified from Columbia University Irving Medical Center, Weill Cornell Medicine, University of Hawaii Cancer Research Center, the Iowa Cancer Registry at the University of Iowa, and the Eastern Division of Cooperative Human Tissue Network (CHTN). Subjects who were found to have occult lymph node metastasis following initial surgery were excluded. Subjects with the Eastern Cooperative Oncology Group (ECOG) performance-status score of 0 (no symptoms) or 1 (mild symptoms) were included.

The following clinico-demographic information was obtained from the electronic clinical record: age at diagnosis, gender, race/ethnicity (white non-Hispanic, white Hispanic, black non-Hispanic, black Hispanic, Asian), TNM stage (I vs II), histologic tumor grade (well vs moderate-to-poorly differentiated), treatment received (surgery with or without neck dissection, irradiation, or both neck dissection and irradiation), tobacco use (never/former vs current) and alcohol abuse (4 or more drinks on any day or 8 or more drinks per week; never/former vs current). Time of initial surgical treatment until cancer recurrence and cancer-specific death were also noted.

For each subject, archived formalin-fixed paraffin-embedded (FFPE) tissue blocks were retrieved. In case the subject had recurrent and/or second primary OSCC, the initial OSCC surgical tissue sample was utilized for the analysis. Ten 10-μm sections were obtained from archived FFPE tumor tissue samples for all subjects. For each sample, a representative section was stained with H&E and reviewed by a pathologist to identify regions containing >90% malignant epithelial cells for microdissection as previously described [7]. Total RNA was isolated from tissues using RNeasy FFPE kits (Qiagen Inc., Valencia, CA) following the manufacturer’s protocol, yield was quantitated by Nanodrop, and samples were stored at −80° C.

### MicroRNA expression assessment by quantitative real-time PCR (qRT-PCR)

The prognostic miRNA-127-3p, 4736, 655-3p expression levels were quantified using the miScript II Rt kit (Qiagen) as described previously [7]. Briefly, 9 µL of isolated RNA was added to the cDNA master mix, composed of 5x miScript HiSpec Buffer, 10x miScript Nucleics Mix, miScript Reverse Transcriptase Mix, and water, to a total volume of 20 µL. The cDNA was incubated at 37 °C for 60 min, followed by 5 min incubation at 95 °C and then diluted 11 times. For amplification reactions, the miScript miRNA PCR Custom Array with a miScript SYBR Green PCR kit (Qiagen) was used in a 7300 qPCR system (Applied Biosystems Inc., Beverly, MA), following the cycling conditions recommended by the supplier (15 min at 95 °C, followed by 40 cycles of 15 s at 94 °C, 30 s at 55 °C, and 30 s at 70 °C). The coefficient of variation was calculated and values <5% were considered acceptable. Test samples were assayed in duplicate with the laboratory blinded to survival status and with 5% duplication after relabeling. Data was analyzed to determine the threshold cycle (Ct). The endogenous control RUN6-6p was used to normalize the relative expression of target miRNAs (ΔCt). The samples with undetermined Ct value for the control were excluded from analysis. Those with an undetermined Ct for specific miRNAs were assigned a value of 39.99.

### Mortality risk score calculation

Using the miRNA-based mortality risk score formula, the risk score was calculated for all subjects as previously described [7]. The mortality risk scores were obtained by summing the expression values of the selected miRNAs and covariates weighted by the regression coefficients obtained from the multivariate Cox regression analyses. **Mortality risk score** = (- 0.7 x expression value of miRNA-127-3p) + (−0.3 x expression value of miRNA-4736) + (0.1 x expression value of miRNA-655-3p) + (0.9 x 0 for TNM Stage I; 1 for TNM Stage II) + (0.4 x 0 for well-differentiated; 1 for moderately/poorly differentiated), in which the miRNA expression level is the ΔCt value of each miRNA. Based on the individual mortality risk score, the patients were first stratified into high (≥0) vs. low (<0) mortality risk groups, and then further stratified into highest (≥2), moderately-high (1 to <2), moderately-low (0 to <1) and low (<0) risk groups.

### Prognostic miRNA functional analysis

Network visualization and functional analysis were previously performed using Cytoscape v3.2.0 to identify potential gene targets of miRNA-127-3p, 4736, 655-3p [7]. We used the gene targets from the previous study to identify the intersection among genes that control cancer recurrence and aggressiveness. Venn diagram was constructed to visualize gene targets common to three miRNAs.

### Statistical analysis

We calculated the individual mortality risk scores for all patients in the four treatment groups using our miRNA-based mortality risk score formula. Based on these mortality risk scores, the patients were stratified into high (≥0) vs. low (<0) mortality risk groups. For each risk group, the median time-to-death and the median time-to-recurrence, as well as the 5-year survival rate was assessed. The hazard ratios (HR) of the treatments S+ND, S+IR, and S+ND/IR were computed over S. The Kaplan-Meier curves were generated for the high and low-risk strata of the four treatment groups. Statistical analyses were conducted using R.

## RESULTS

### Subject characteristics

The demographic and clinico-pathologic characteristics of each treatment category is shown in **Table 1** (total n=836). The treatment groups include surgery only (S; n=551), surgery with neck dissection (S+ND; n=164), surgery with radiotherapy (S+IR; n=76) and surgery with neck dissection and irradiation (S+ND/IR; n=45). Compared to S group, significantly more patients with higher TNM stage (Stage II) and histologic tumor grade (moderately/poorly-differentiated carcinoma) received treatment in addition to surgery.

**Table 1.** Clinical and histopathologic characteristics of the early-stage oral squamous cell carcinoma patients included in this study.

|  | **S Only** | **S+ND** | **S+IR** | **S+ND/IR** |
| --- | --- | --- | --- | --- |
| **Patients** | n=551 | n=164 | n=76 | n=45 |
| **Age** |  |  |  |  |
| Mean (range) | 66 (21-97) | 61 (25-87) | 65 (24-90) | 58 (35-80) |
| **Gender** |  |  |  |  |
| Female | 218 (40%) | 84 (51%) | 24 (32%) | 21 (47%) |
| Male | 333 (60%) | 80 (49%) | 52 (68%) | 24 (53%) |
| **Race/Ethnicity** |  |  |  |  |
| White non-Hispanic | 383 (70%) | 92 (56%) | 30 (39%) | 22 (49%) |
| White Hispanic | 80 (15%) | 15 (9%) | 9 (12%) | 9 (20%) |
| Black non-Hispanic | 22 (4%) | 4 (2%) | 1 (1%) | 7 (6%) |
| Black Hispanic | 5 (1%) | 0 (0%) | 0 (0%) | 0 (0%) |
| Asian | 52 (9%) | 34 (21%) | 27 (36%) | 5 (11%) |
| Unknown | 9 (2%) | 19 (12%) | 9 (12%) | 2 (4%) |
| **TNM Stage** |  |  |  |  |
| Stage I | 428 (78%) | 81 (49%) | 40 (53%) | 17 (38%) |
| Stage II | 123 (22%) | 83 (51%) | 36 (47%) | 28 (62%) |
| **Histologic Grading** |  |  |  |  |
| Well-differentiated | 291 (53%) | 76 (46%) | 29 (38%) | 8 (18%) |
| Moderately/Poorly-differentiated | 260 (47%) | 88 (54%) | 47 (62%) | 37 (82%) |
| **Smoking Status** |  |  |  |  |
| Never | 163 (30%) | 64 (39%) | 2 (3%) | 17 (38%) |
| Past | 72 (13%) | 39 (24%) | 9 (12%) | 7 (16%) |
| Current | 54 (48%) | 12 (7%) | 5 (7%) | 6 (13%) |
| Unknown | 262 (48%) | 49 (30%) | 60 (79%) | 15 (33%) |
| **Alcohol Abuse** |  |  |  |  |
| Never | 201 (36%) | 80 (49%) | 4 (5%) | 22 (49%) |
| Past | 32 (6%) | 14 (9%) | 6 (8%) | 0 (0%) |
| Current | 56 (10%) | 21 (13%) | 6 (8%) | 8 (18%) |
| Unknown | 262 (48%) | 49 (30%) | 60 (79%) | 15 (33%) |

### Mortality risk score-based stratification

The miRNA-based mortality risk score formula was utilized to calculate the risk score for all patients. Within each treatment group, the patients were stratified into either high-risk (≥0) vs low-risk (<0). The median time-to-recurrence and time-to-death, as well as 5-year survival rates were calculated for high vs. low risk strata in each treatment group (**Table 2**). Within the high-risk strata, S+ND significantly improved the 5-year survival (75%) over S (46%). Compared to S+ND, S+IR was not associated with survival benefit (44% 5-year survival rate for S+IR and 62% for S+ND/IR) as shown in **Figure 1a**. The HR of S+ND over S was 0.37 (95% CI: 0.2-0.6) with a p-value of 0.0005, indicating survival benefit with S+ND for high-risk patients.

**Table 2.** The efficacy of various treatment regimens in oral squamous cell carcinoma patients stratified into high (risk score ≥0) versus low-risk (risk score <0) using the microRNA-based prognostic model; surgery (S) only, surgery with neck dissection (S+ND), surgery with irradiation (S+IR) and surgery with neck dissection and irradiation (S+ND/IR).

| Treatment | Risk Category | Median Time-to-Recurrence | Median Time-to-Death | Cancer Death | 5-year Survival |
| --- | --- | --- | --- | --- | --- |
| **S** | High risk ($>0$) | 26 (months) | 50 (months) | 54% | 46% |
| | Low risk ($<0$) | $\geq 60$ | $\geq 60$ | 11% | 89% |
| **S + ND** | High risk | $\geq 60$ | $\geq 60$ | 25% | 75%* |
| | Low risk | $\geq 60$ | $\geq 60$ | 26% | 74%* |
| **S + IR** | High risk | 41 | 43 | 56% | 44% |
| | Low risk | 33 | $\geq 60$ | 49% | 51%* |
| **S + ND/IR** | High risk | 7 | $\geq 60$ | 38% | 62% |
| | Low risk | $\geq 60$ | $\geq 60$ | 34% | 66%* |

\* $p \leq 0.05$ when compared with the same risk strata of S only.

**Figure 1a.**
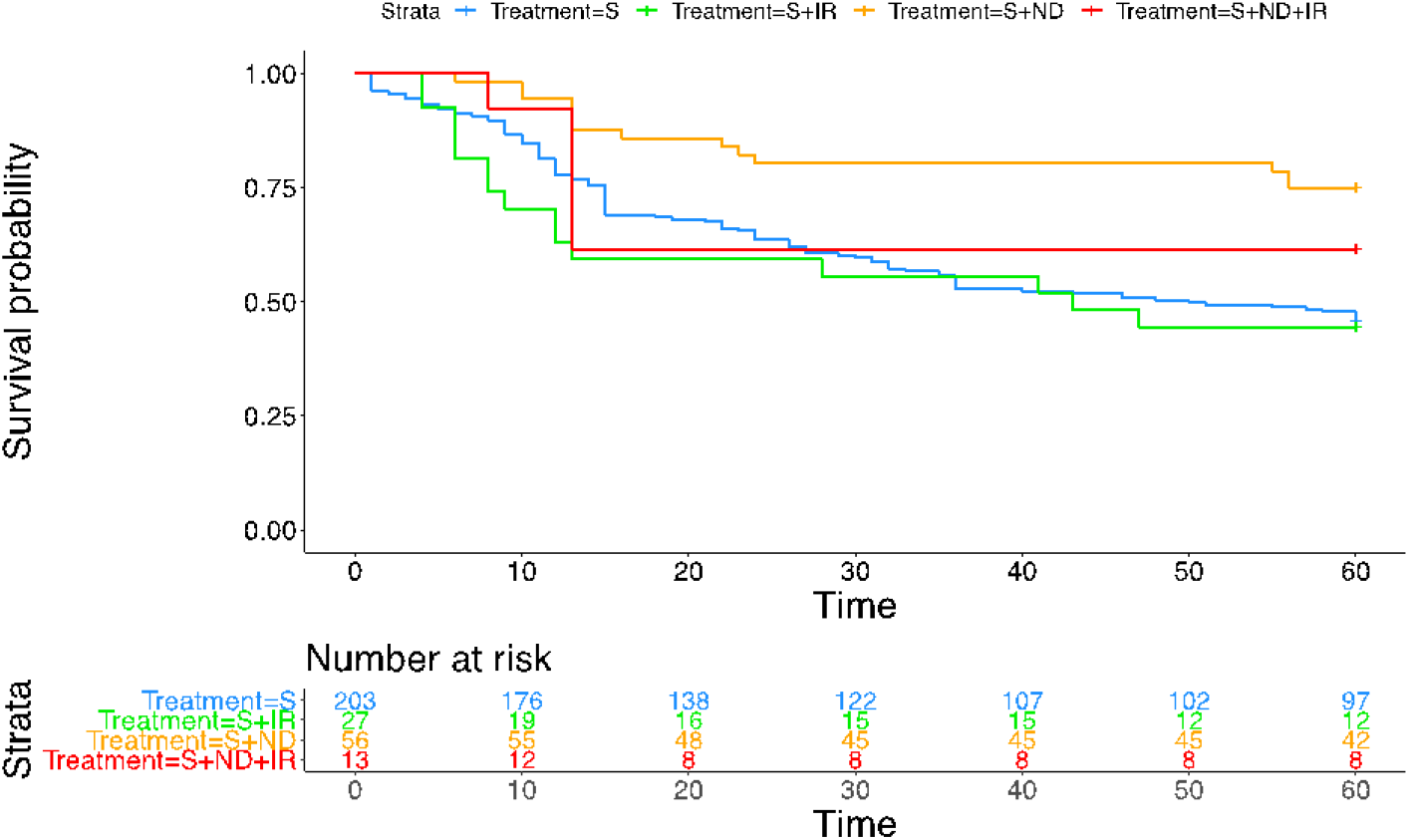
Kaplan-Meier curve demonstrating survival benefit of surgery with neck dissection over other treatment modalities in oral squamous cell carcinoma patients with high mortality risk scores (≥0). *Time in months.

In the low-risk strata, S was associated with the highest 5-year survival rate compared to other treatment modalities (Table 2). S+ND that demonstrated significant survival benefit in the high-risk strata, was associated with lower 5-year survival of 74% compared to S (5-year survival rate of 89%) in the low-risk strata. Similarly, S+IR and S+ND/IR did not offer survival benefit over S for low-risk patients. The HR of S+ND over S was 2.6 (95% CI: 1.6-4.2), for S+IR 6.5 (95% CI: 3.9-10.8) and for S+ ND/IR 3.4 (95% CI: 1.7-6.7) all with p-values ≤0.0001. Thus, for the low mortality risk strata, surgery only was the best treatment option (**Figure 1b**).

**Figure 1b.**
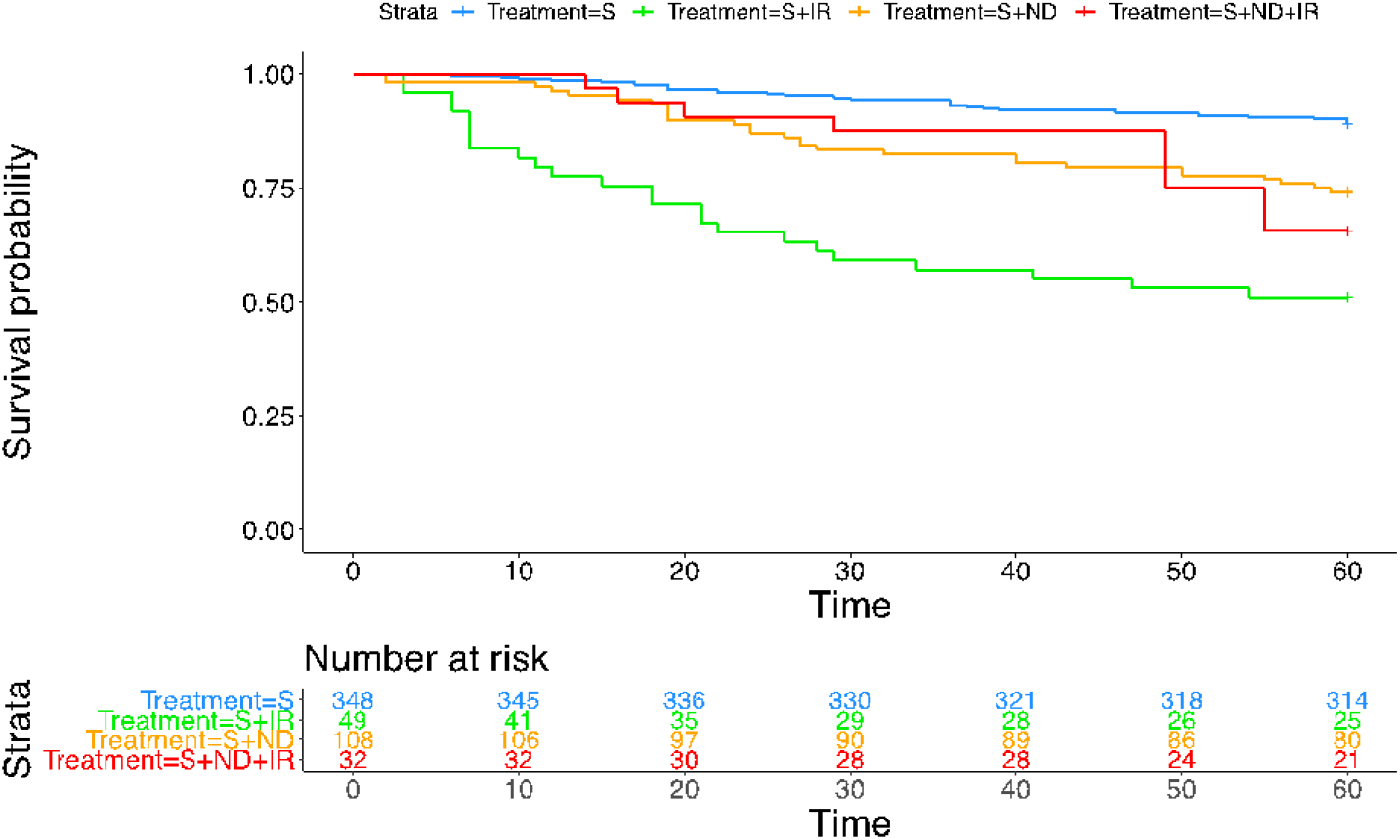
Kaplan-Meier curve delineating survival benefit of surgery only over other treatment modalities in oral squamous cell carcinoma patients with low mortality risk scores (<0). *Time in months.

### Sub-risk strata analysis of S versus S+ND

For the patients who were treated with S or S+ND, we compared the median time-to-recurrence, the median time-to-death and 5-year survival rate by sub-stratifying risk categories to highest (≥2), moderately-high (<2-1), moderately-low (<1-0) and low (<0) as shown in **Table 3**. For the patients in the moderately-high and moderately-low sub-strata, the 5-year survival rates were 29% and 58%, respectively with S. However, if these patients received S+ND instead, the 5-year survival rate increased to 100%. The patients with risk score ≥2 who had S as initial treatment demonstrated significantly shorter median time-to-recurrence (6 months), median time-to-death (11 months), and the 5-year survival rate of 6%; With S+ND, the survival rate improved to 68%. For the patients in the low-risk sub-strata (risk score <0), S consistently demonstrated survival benefit over S+ND. The clinical risk categories, high (risk score ≥2) vs. moderate (risk score between 0 to <1) vs. low risk (risk score <0), can be used as a guideline to select initial surgical treatment and also to identify high-risk patients (risk score ≥2) who may need therapy additional to surgery to optimize survival.

**Table 3.** The median time-to-recurrence, time-to-death and survival rate in oral squamous cell carcinoma patients treated with surgery only (S) or surgery with neck dissection (S+ND). Initial high versus low risk stratification using 0 as cutoff, and then sub-risk stratification into highest, moderately-high, moderately-low and low risks are shown. For clinical use, the patients can be categorized into high versus moderate versus low risk groups; the ‘high-risk strata (≥2)’ represents a group of patients at greater risk of cancer relapse and death even after S+ND, compared to ‘moderate-risk strata (0 to <2)’, in which survival greatly improves with S+ND. For the ‘low-risk strata (<0)’, S is associated with survival benefit.

| Risk Strata | Sub-Risk Strata | Risk Scores | Median Time-to-Recurrence | Median Time-to-Death | Cancer Death | 5-year Survival | Clinical Risks |
| --- | --- | --- | --- | --- | --- | --- | --- |
| High-risk | **Highest risk** | $\geq 2$ | | | | | High-risk |
|  | S only |  | 6 (months) | 11 (months) | 94% | 6% |  |
| | S+ND | | 58 | $\geq 60$ | 32% | 68% | |
| | **Moderately-high risk** | 1 to $<2$ | | | | | Moderate-risk |
|  | S only |  | 14 | 22 | 71% | 29% |  |
| | S+ND | | $\geq 60$ | $\geq 60$ | 0% | 100% | |
| | **Moderately-low risk** | 0 to $<1$ | | | | | |
| | S only | | 46 | $\geq 60$ | 42% | 58% | |
| | S+ND | | $\geq 60$ | $\geq 60$ | 0% | 100% | |
| Low-risk | **Low risk** | $<0$ | | | | | Low-risk |
| | S only | | $\geq 60$ | $\geq 60$ | 11% | 89% | |
| | S+ND | | $\geq 60$ | $\geq 60$ | 26% | 74% | |

### Functional analysis of prognostic miRNAs

We had previous identified common gene targets of miRNA-127-3p, 4736, 655-3p [7]. In this study, we assessed the intersection among these genes that control cancer recurrence and aggressiveness (**Figure 2**). The Rac family small GTPase 1 (Rac1) and the Rho GTPases activated p21-activated kinases (PAKs) pathway, including Cdc42 and hTERT were identified as key targets of the miRNAs. Rho GTPases are crucial regulators of cell migration and are altered in many cancer types, including colon, glioblastoma and head and neck cancer [15]. Rac, Rho and Cdc42 are subfamilies of Rho GTPase [16]. Integrin-mediated cell-extracellular matrix adhesion activates Rac1, which directly binds and activates PAKs and other effectors including phosphatidylinositol-4-phosphate 5-kinase, Nap125, PIR121 and IRSp53, resulting in increased membrane protrusions and actin polymerization [16,17]. The other member of Rho GTPase subfamily Cdc42, once activated, phosphorylates PAK1 and PAK2, which in turn leads to filopodia formation [17]. These changes either by themselves or integrated can affect gene expression, cell cycle progression and apoptosis [17]. Importantly, Cdc42/Rac1 participates in the post-transcriptional control of telomerase activity by transcriptional upregulation of human telomerase reverse transcriptase (hTERT) [18,19]. Hence, PAKs/Rac1-Cdc42-hTERT signaling pathway may play a central role in cancer recurrence and aggressiveness.

**Figure 2.**
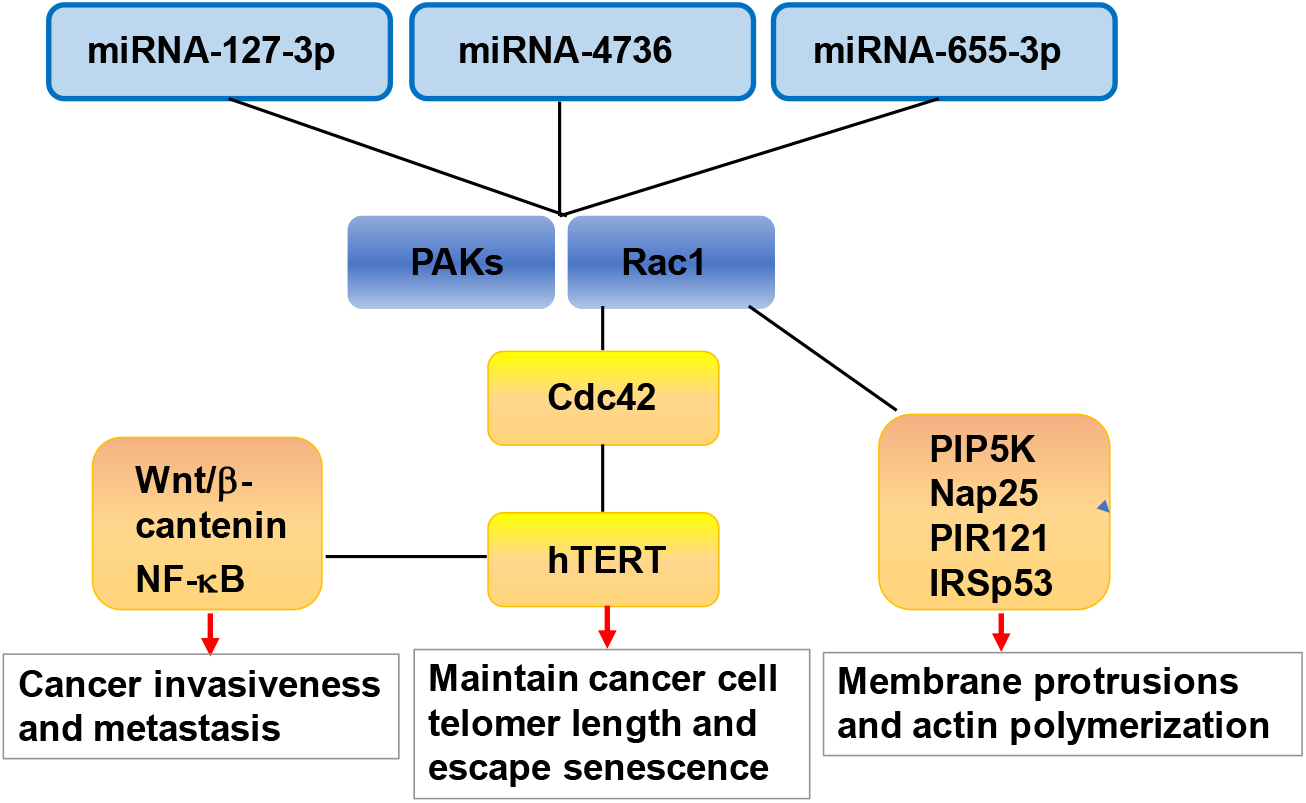
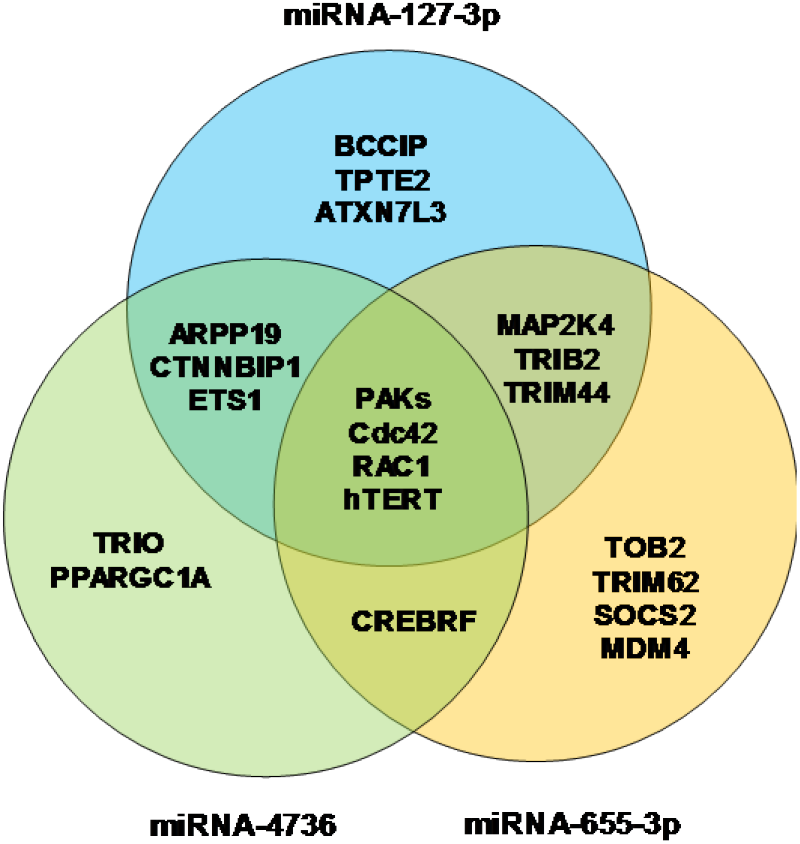
Common gene targets of microRNA-127-3p, 4736, 655-3p that are associated with cancer recurrence and aggressiveness. **a.** The cancer-related pathway involving Rac family small GTPase 1 (Rac1), p21-activated kinases (PAKs), Cdc42, and human telomerase reverse transcriptase (hTERT) was targeted by three microRNAs. **b.** Venn diagram showing common gene targets of three microRNAs. **Figure 2a.** Gene targets. **Figure 2b.** Venn diagram.

### Expression level analysis of Rac1, Cdc42 and hTERT by qRT-PCR

We assessed expression levels of Rac1, Cdc42 and hTERT using qRT-PCR. Due to the shortage of samples and materials, the expression levels of three gene targets were studied in forty one patients who had S or S+ND. Gene expression levels were also assessed in oral tissue samples from five healthy individuals for comparison. cDNA was made from total RNA using high-capacity cDNA Reverse Transcription Kit (Applied Biosystems, USA) according to the manufacturer’s protocol. The expression levels were measured by qRT-PCR (LightCycler 480 II, Roche Applied Science). The following TaqMan Gene Expression Assays were used (Applied Biosystems): hTERT (Hs00972650_m1), Cdc42 (Hs0198044_g1), and Rac1 (Hs01902432_s1). The reactions were carried out in a total volume of 12.5 μl, with 1X power TaqMan Fast Advanced Master Mix and 0.2 μM of each primer. Data was analyzed to determine the threshold cycle (Ct). The expression levels were normalized using GAPDH as an endogenous control (ΔCt). The ΔΔCt and fold change in expression levels calculated for each sub-risk strata compared to the healthy controls are shown in **Table 4**.

**Table 4.** The ΔΔCt and the fold change in the expression levels of Rac1, Cdc42, and hTERT in four sub-risk strata compared to healthy controls. The hTERT expression levels significantly correlated with the risk scores.

| Sub-Risk Strata | Risk Score | $\Delta\Delta\text{Ct}$ | | | Fold Change | | |
| --- | --- | --- | --- | --- | --- | --- | --- |
|  |  | Rac1 | Cdc42 | hTERT | Rac1 | Cdc42 | hTERT |
| Highest Risk | $\geq 2$ | 3.22 | 1.05 | 4.55 | 8.32 | 1.07 | 22.43* |
| Moderately-High Risk | 1 to <2 | 2.95 | 0.88 | 3.86 | 6.71 | 0.84 | 13.53* |
| Moderately-Low Risk | 0 to <1 | 2.77 | 0.23 | 3.61 | 5.81 | 0.17 | 11.17* |
| Low Risk | <0 | 2.00 | 0.21 | 3.42 | 3.01 | 0.16 | 9.78* |
| Healthy Controls | - | Reference | Reference | Reference | Reference | Reference | Reference |

\* $p \leq 0.05$ when compared with the healthy controls.

hTERT expression levels were mostly undetectable in the control samples, and Ct of 39.99 was used. hTERT demonstrated 22-fold increase in the highest-risk sub-strata compared to healthy controls (p<0.0005). Rac1 and Cdc42 were also overexpressed in the highest-risk sub-strata, although statistical significance was not reached. The expression levels of three gene targets correlated with the risk scores, demonstrating that elevated hTERT, Rac1, Cdc42 levels are associated with cancer recurrence and poor prognosis.

### hTERT expression by quantitative immunofluorescent assay

To further assesse hTERT expression in more aggressive form of OSCC, we performed quantitative immunofluorescent assays in ten patients with risk scores of 2 or higher who had cancer recurrence. The formalin-fixed paraffin-embedded tissue slides were stained with anti-human hTERT (Rockland Immunochemicals, Limerick, PA). The hTERT stain concentration and the number of nuclei staining with hTERT were quantified using a macro derived from the Leica Quantitative Algorithm v1. The hTERT stain concentration was multiplied by the number of hTERT-expressing tumor cells to obtain final hTERT expression levels. After adjusting for the outliers, we had eight evaluable cases. Differential hTERT expression levels were compared between those who had cancer-specific death following recurrence (n=4) vs those who had 5-year survival despite having a recurrence (n=4) (**Figure 3a**). hTERT levels were 2.4-fold higher in those who had a cancer-specific death (an average hTERT expression level of 8,061; an average hTERT stain concentration of 3.8, with an average of 2,318 hTERT-expressing tumor cells) compared to those with 5-year survival (an average hTERT expression level of 3,330; an average hTERT stain concentration of 3.5, with an average of 884 hTERT-expressing tumor cells). hTERT level correlated with the mortality risk scores, demonstrating that elevated hTERT level is associated with poor survival.

**Figure 3.**
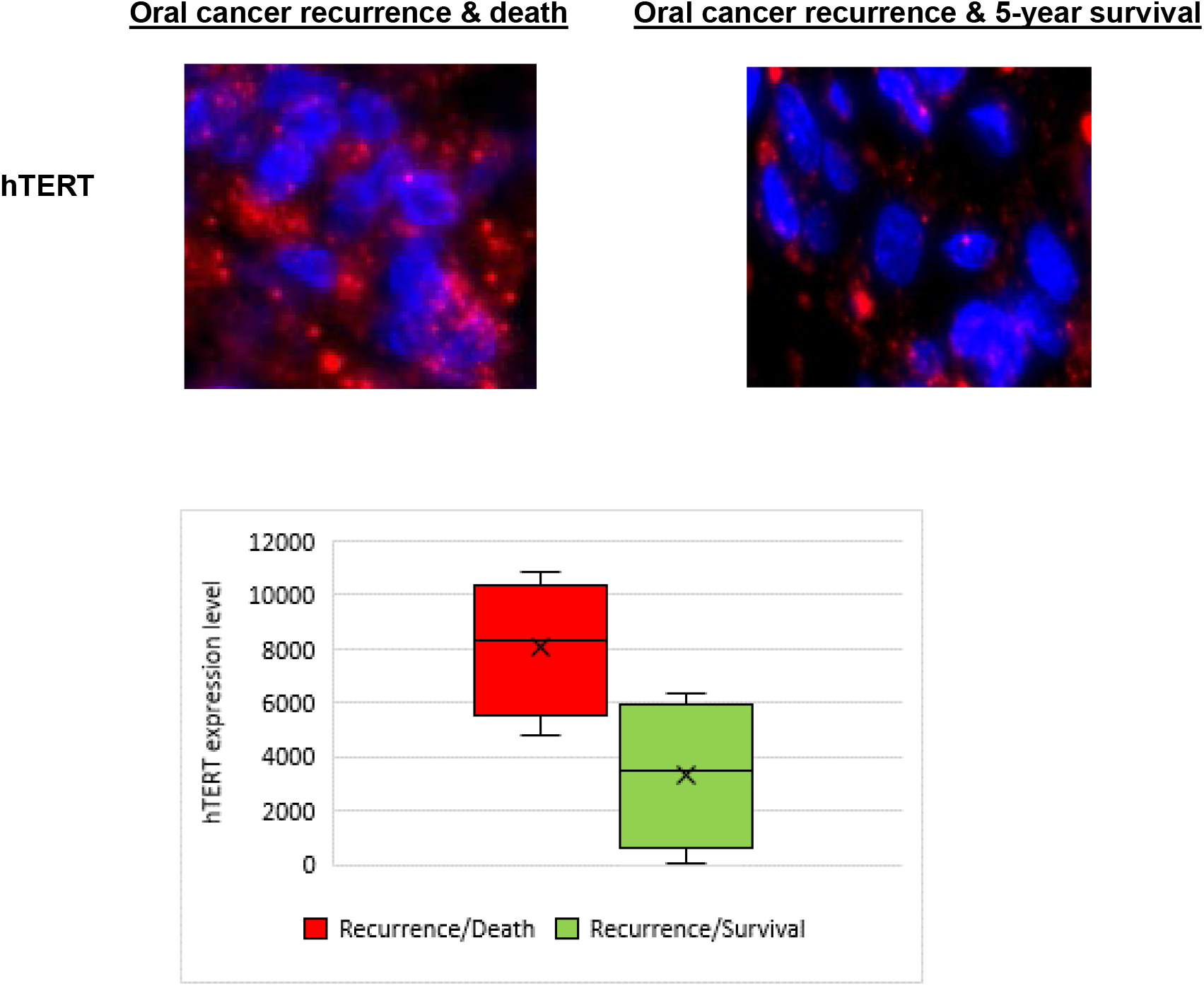
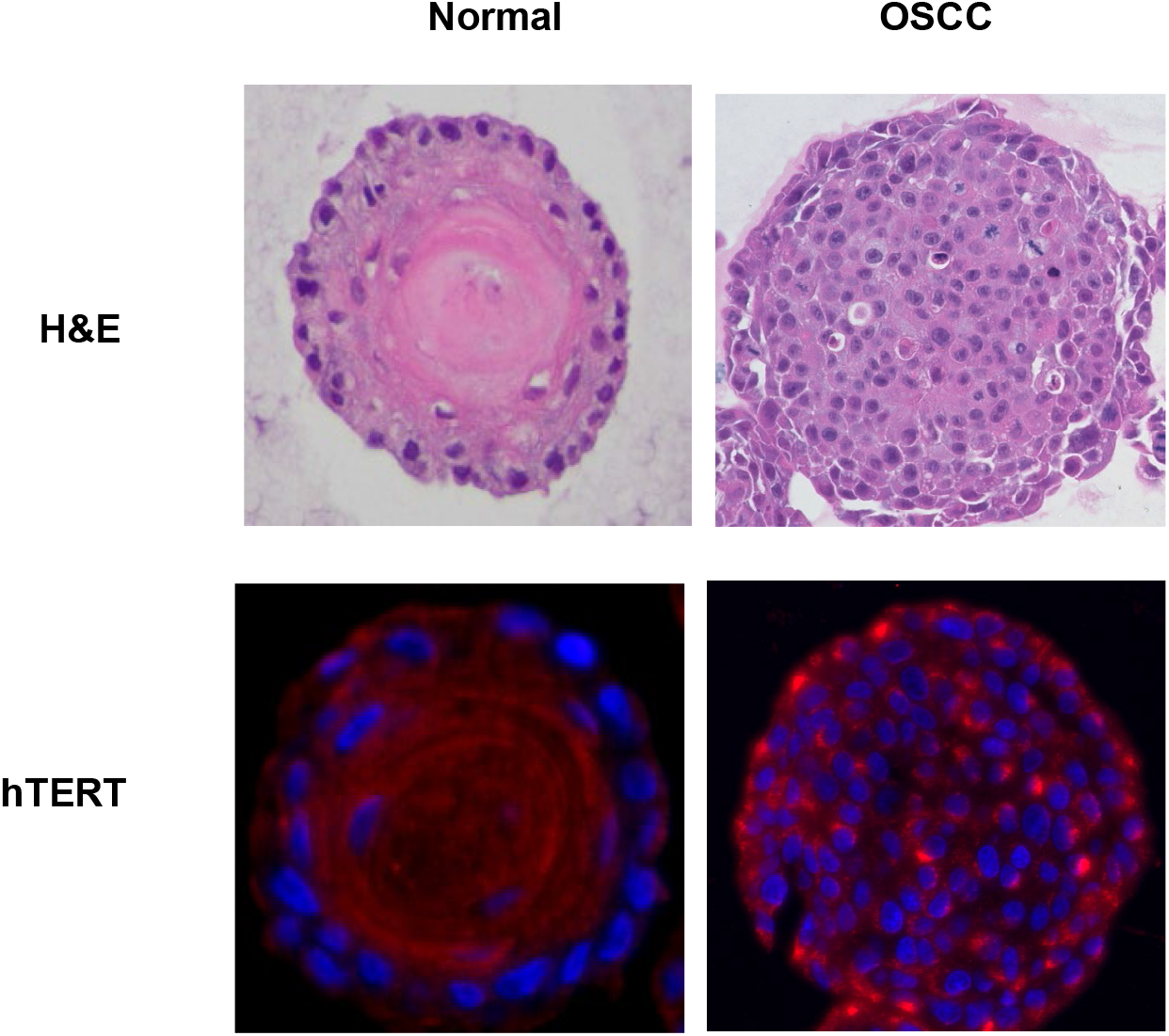
The hTERT expression levels assessed by quantitative immunofluorescence assay. **Figure 3a.** Elevated hTERT expression levels (bright red dots on blue DAPI-stained nuclei) were observed in patients with poor prognosis (cancer recurrence and death within 5-year period) compared to those who survived the 5-year period despite having cancer recurrence. Dot plots comparing hTERT expression levels in poor versus favorable prognostic group. Bars indicate the mean hTERT expression level. **Figure 3b.** Minimal hTERT expression level was detected in normal oral organoids. In comparison, elevated hTERT expression level (bright red nuclear and peri-nuclear dots) was observed in oral squamous cell carcinoma (OSCC) organoids.

We also assessed feasibility of hTERT level quantification in the patient-derived normal and OSCC organoids. We first obtained small piece (5×5×3mm) of normal and OSCC tissue from patients at the time of surgery. Surface epithelium from normal tissues and tumor islands within lamina propria from OSCC tissues were isolated. Tissue sections were minced, enzymatically dissociated, and forced through a 70 μm cell strainer. The single cell suspension (4×10^5^/ml) was mixed with Matrigel (Corning, Corning, NY) and 2×10^4^ cells in 50 μl of Matrigel were seeded into the 24-well plates. Organoids were grown for 10-14 days at 37°C in a humidified atmosphere of 5% CO_2_ as previously described [19]. Under an inverted phase-contrast microscope, growing organoids were observed and photomicrographed to determine their number and size. Organoid formation rate was defined as the average number of ≥50 μm spherical structures at day 14 divided by the total number of cells seeded in each well at day 0. Organoids were recovered and fixed overnight in paraformaldehyde for histological examination and hTERT quantitative immunofluorescent assay. There were minimal hTERT immunofluorescent staining in normal organoids and stronger immunofluorescent staining in OSCC organoids (**Figure 3b**).

## DISCUSSION

Currently there is no clinical modality to identify patients at high-risk for cancer-specific death among those assigned to early-stage OSCC (TNM stages I & II). Because 80% of oral cancer patients are in early-stage at the time of diagnosis [2], a window of opportunity exists in which proper prognostication and subsequent decisions for additional treatment can dramatically improve the 5-year survival of patients with this deadly disease.

Using the miRNA-based mortality risk score formula, we assigned a risk score to all patients and assessed the survival benefit in high (≥0) vs. low (<0) risk strata based on the treatment received. Compared to S, S+ND significantly increased the median time-to-recurrence from 26 months to ≥60 months and the 5-year survival rate from 46% to 75% (p<0.001) for the high-risk group, consistent with other reports that S+ND is associated with survival benefit in early-stage OSCC [20,21]. Improved 5-year survival in the S+ND group was most likely secondary to prolonged time-to-recurrence. Indeed, initial treatment modality and time-to-recurrence were reported to be independent prognostic variables [22].

Paradoxically, the patients in the low mortality risk group who received S+ND had a lower 5-year survival rate of 74% compared to 89% in the S only group. The significant reduction in the 5-year survival in the low-risk strata who had S+ND may be explained by the low success rate of salvage surgery at the time of cancer recurrence [22,23]. With initial S+ND treatment, patients experienced recurrence in a less predictable fashion, often involving the contralateral neck [22,23]. In contrast, patients treated with S only were more likely to present with nodal disease confined to the neck at levels I, II, and III, which were successfully removed during salvage surgery [22,23]. Consistently, the high-risk patients in our study who were initially treated with S+ND, 54% experienced cancer recurrence and died of disease due to failed salvage surgery. S+IR and S+IR+ND demonstrated minimal survival benefit over S or S+ND for both high and low-risk strata.

Based on the assessment of sub-risk strata [highest (≥2), moderately-high (<2-1), moderately-low (<1-0) and low (<0)], it may be beneficial for patients to be risk stratified using the prognostic marker at the time of biopsy. For those with low mortality risk scores (<0; low clinical risk), surgery only without neck dissection is the most beneficial treatment modality. On the other hand, for those with risk scores between 0 and ≤1 (moderate clinical risk), S+ND instead of S will improve the 5-year survival rate to 100%. However, for the patients with the highest risk score of ≥2 (high clinical risk), while S+ND improves the 5-year survival rate over S to 68%, it is still suboptimal. These patients may benefit from additional therapy following surgery to prevent cancer recurrence, which will further improve the 5-survival rate.

Human telomerase reverse transcriptase (hTERT) along with few other molecules in the same signaling pathway were demonstrated to be targeted by all three miRNAs. Indeed, there was 22-fold increase in the hTERT expression levels in the highest-risk sub-strata compared to healthy controls (p<0.0005). Consistently, our quantitative immunofluorescence study demonstrated elevated levels of hTERT in OSCC patients who had cancer recurrence and death, compared to those who survived despite having cancer relapse. In patient-derived organoids, we showed that there is overexpression of hTERT in OSCC and minimal expression in normal organoids. Others have reported similar findings. Elevated expression of hTERT was associated with a poor prognosis in solid tumors such as gastric, lung, cervical, head and neck, breast, ovarian cancer, and glioblastoma [24,25]. In head and neck cancer patients, elevated hTERT expression was associated with a higher recurrence rate (p = 0.044) and a lower 5-year survival rate (p = 0.011) [26]. An elevated level of hTERT was also observed in oral cells within the cancerized field [27]. Moreover, hTERT expression level correlated with degree of oral preneoplasia [27]. Compared to normal oral mucosa, hTERT expression was elevated by a 6.9-fold in OSCC [28]. Thus, the hTERT signaling axis is a viable therapeutic target for oral cancer patients.

Three-dimensional (3D) tumor organoid models have been consistently shown to faithfully recapitulate features of the tumor of origin in terms of cell differentiation, heterogeneity, histoarchitecture and clinical drug response [29-32]. Thus, there is increasing interest in developing tumor organoid models for drug development and personalized medicine applications. Functional precision therapy approaches where the primary tumor tissue is directly exposed to drugs to determine efficacy have the potential to boost personalized medicine efforts and influence clinical decisions [33-36]. A recent study found that patient-derived organoids could accurately predict patient responses to therapy, with 100% sensitivity and 93% specificity [37]. While establishing patient-derived xenografts is costly and time-consuming, *in vitro* 3D organoids derived from primary cancers can be established rapidly, with successful passage within 10-14 days of *in vitro* growth and with >80% efficiency [29]. Thus far, we have cultivated patient-derived organoids from normal oral epithelium and OSCC. These organoids can be utilized in the future as a functional *in vitro* testing platform to explore novel therapeutic options, which has a potential role in clinical decision-making tailored to each individual.

## CONCLUSION

In response to the critical need to subdivide traditional tumor classes into subsets that behave differently and also to refine and improve prognostication and treatment selection, we have developed and validated a novel miRNA-based prognostic model to predict survival outcome in patients who are already categorized into ‘early-stage’ by the TNM system. For clinical practicality, we developed a parsimonious risk score formula that is capable of translating miRNA expression levels assessed by qRT-PCR at the time of initial cancer diagnosis into a score that reflects the risk of cancer-specific mortality during the 5-year period from the time of surgery. Improved survival was observed in the high-risk strata who received S+ND and in the low-risk strata receiving S as the initial treatment. Moreover, we demonstrated that for patients with risk scores of ≥2, there is suboptimal increase of 5-year survival to 68% with S+ND. Hence, these patients may benefit from therapeutic intervention in addition to surgical treatment to further improve survival. We also demonstrated that hTERT overexpression is associated with shorter median time-to-recurrence and death, making it a promising drug target. Cancer vaccines that prime immune system against hTERT-expressing cells are currently undergoing clinical trials for various solid tumors [38-40]. If delivered following surgical treatment, these vaccines can be repurposed to prevent cancer recurrence and improve survival in high-risk early-stage oral cancer patients.

## Data Availability

The data provided in this publication will be available from the corresponding author upon request.

## Data Availability

The data provided in this publication will be available from the corresponding author upon request.

## Acknowledgements

We thank Charis Yoon for her assistance in data organization, and Qiao Wang for her technical assistance.

